# Molecular Surveillance of Antimicrobial Resistance in Human Clinical Isolates: A Clinician–Scientist Growth Journey from Rural Gujarat, India

**DOI:** 10.64898/2025.12.27.25343078

**Authors:** Atul Amarshibhai Devganiya

**Affiliations:** Department of Community Medicine, Kiran Medical College, Surat, Gujarat, India; Formerly: Banas Medical College & Research Institute, Palanpur, Gujarat, India

**Keywords:** Antimicrobial resistance, ESBL, carbapenemase, MRSA, VRE, molecular surveillance, rural India

## Abstract

**Background:** Antimicrobial resistance (AMR) represents one of the most serious threats to global public health, with a disproportionate burden borne by low- and middle-income countries. Rural healthcare settings in India remain under-represented in molecular AMR surveillance despite high antimicrobial use and limited diagnostic capacity.

**Objectives:** To describe longitudinal phenotypic and molecular AMR patterns in clinical bacterial isolates from a rural district hospital in Gujarat, India, and to assess the feasibility of clinician-led molecular surveillance in a resource-limited setting.

**Methods:** A retrospective observational study was conducted on 242 non-duplicate discarded clinical bacterial isolates collected between January 2021 and August 2025 at Banas Civil Hospital, Palanpur, Gujarat. Antimicrobial susceptibility testing was performed according to Clinical and Laboratory Standards Institute guidelines (CLSI M100, 31st edition) using Kirby–Bauer disk diffusion and broth microdilution methods. Molecular detection of blaCTX-M, blaNDM, blaOXA-48, mecA, and vanA resistance genes was carried out using polymerase chain reaction, with confirmation by Sanger sequencing. Data were analysed using R software (version 4.3.2).

**Results:** The most common pathogens were *Escherichia coli* (38%), *Klebsiella pneumoniae* (26%), and *Pseudomonas aeruginosa* (17%). Resistance to third-generation cephalosporins was high in *E. coli* (64%) and *K. pneumoniae* (59%). Carbapenem resistance peaked at 17% in 2024. Molecular analysis identified blaCTX-M in 42% of Gram-negative isolates, while blaNDM and blaOXA-48 were detected in 11% and 7%, respectively. Genotype–phenotype concordance exceeded 80%.

**Ethical Approval:** Ethical approval for the study was obtained from the Institutional Ethics Committee of Banas Medical College and Research Institute, Palanpur, Gujarat, India. No patient identifiers were accessed, and no patient interviews, direct contact, or additional specimen collection were performed for research purposes.

**Conclusion:** This study demonstrates a substantial burden of ESBL- and carbapenemase-mediated antimicrobial resistance in rural Gujarat. Clinician-led molecular surveillance using discarded clinical samples is feasible and essential for guiding empirical therapy and strengthening antimicrobial stewardship in resource-limited settings

## 1. Introduction

Antimicrobial resistance (AMR) has emerged as a defining challenge of modern medicine, undermining decades of progress in infection control and therapeutic innovation. Globally, AMR is estimated to contribute to nearly five million deaths annually, with projections indicating further escalation in the absence of effective containment strategies. India represents a major epicentre of this crisis due to high antibiotic consumption, over-the-counter availability, and heterogeneous infection control practices.

National surveillance initiatives such as the Indian Council of Medical Research (ICMR) AMR Surveillance Network have improved data availability from tertiary centres. However, rural district hospitals-where empirical antibiotic use is common and laboratory infrastructure is limited-remain under-represented in molecular AMR datasets. This gap impairs the formulation of locally appropriate empirical therapy guidelines and delays recognition of emerging resistance mechanisms.

This study reports longitudinal phenotypic and molecular AMR trends in clinical isolates collected over five years from a rural district hospital in Banaskantha, Gujarat. In parallel, it documents the evolution of a clinician-scientist–led surveillance model conducted within routine diagnostic workflows using discarded clinical samples.

## 2. Materials and Methods

### 2.1 Study design and setting

This retrospective observational study was conducted at Banas Civil Hospital, Palanpur, Banaskantha district, Gujarat, India. The hospital is a government-run district healthcare facility serving a predominantly rural population. Microbiological analyses were performed within routine diagnostic laboratories affiliated with Banas Medical College and Research Institute, Palanpur, and subsequently Kiran Medical College, Surat. The study period extended from January 2021 to August 2025.

### 2.2 Inclusion and exclusion criteria

Clinical bacterial isolates recovered during routine diagnostic testing were eligible for inclusion. Only non-duplicate isolates were analysed, defined as the first isolate of a given bacterial species obtained from a single patient per specimen type within a single clinical episode. Duplicate isolates, environmental contaminants, and samples lacking definitive bacterial growth were excluded.

### 2.3 Clinical specimens

Isolates were obtained from routinely processed clinical specimens, including urine, blood, sputum or endotracheal aspirates, wound or pus samples, cerebrospinal fluid, and other sterile body fluids. All specimens were collected as part of standard patient care and processed according to hospital microbiology protocols.

### 2.4 Bacterial identification

Initial bacterial identification was performed using standard microbiological techniques, including Gram staining, assessment of colony morphology, and conventional biochemical tests such as catalase, coagulase, oxidase, indole, citrate, urease, and triple sugar iron agar reactions. From 2023 onwards, selected isolates were additionally confirmed using automated identification systems where available. Identification procedures were consistent with accepted clinical microbiology standards.

### 2.5 Antimicrobial susceptibility testing

Antimicrobial susceptibility testing was performed using the Kirby–Bauer disk diffusion method on Mueller–Hinton agar. Zone diameters were interpreted in accordance with Clinical and Laboratory Standards Institute guidelines (CLSI M100, 31st edition). For isolates suspected of carbapenem resistance, minimum inhibitory concentrations were determined using broth microdilution methods.

Quality control was ensured through the routine use of standard reference strains, including Escherichia coli ATCC 25922, Pseudomonas aeruginosa ATCC 27853, and Staphylococcus aureus ATCC 25923. Results were categorised as susceptible, intermediate, or resistant based on CLSI breakpoints applicable during the study period.

### 2.6 Molecular detection of antimicrobial resistance genes

Molecular analysis was conducted on a subset of isolates collected between 2022 and 2025. Bacterial DNA was extracted using standard extraction protocols. Polymerase chain reaction assays were used to detect selected antimicrobial resistance genes, including blaCTX-M, blaNDM, blaOXA-48, mecA, and vanA, using primers previously described in peer-reviewed literature.

Positive amplification products were confirmed by Sanger sequencing to verify gene identity.

### 2.7 Data management and statistical analysis

Laboratory and antimicrobial susceptibility data were recorded in structured datasets. Descriptive statistical analyses were performed using R software (version 4.3.2). Resistance proportions were calculated for major pathogen–antibiotic combinations. Genotype–phenotype agreement was assessed using Cohen’s kappa statistics. Results are presented as frequencies and percentages.

## 3. Results

### 3.1 Distribution of isolates by year and specimen typeTable 1

presents the year-wise distribution of isolates by specimen type. Blood (38%) and urine (31%) samples predominated throughout the study period, reflecting a high burden of bloodstream and urinary tract infections.

**Table 1.** Distribution of clinical isolates by year and specimen type.

| Year | Total | Urine | Blood | Sputum/ETA | Wound/Pus | CSF/Sterile | Other |
| --- | --- | --- | --- | --- | --- | --- | --- |
| 2021 | 64 | 19 | 31 | 6 | 5 | 3 | 0 |
| 2022 | 87 | 26 | 37 | 8 | 10 | 6 | 1 |
| 2023 | 51 | 24 | 9 | 5 | 4 | 7 | 2 |
| 2024 | 29 | 4 | 9 | 8 | 4 | 3 | 1 |
| 2025 | 11 | 3 | 5 | 2 | 1 | 0 | 0 |
| **Total** | **242** | **76** | **91** | **29** | **24** | **19** | **3** |

### 3.2 Bacterial species distribution

**Table 2** summarises bacterial species distribution. *E. coli* and *K. pneumoniae* together accounted for 64% of all isolates, underscoring their dominant role in both community-onset and hospital-acquired infections.

**Table 2.**
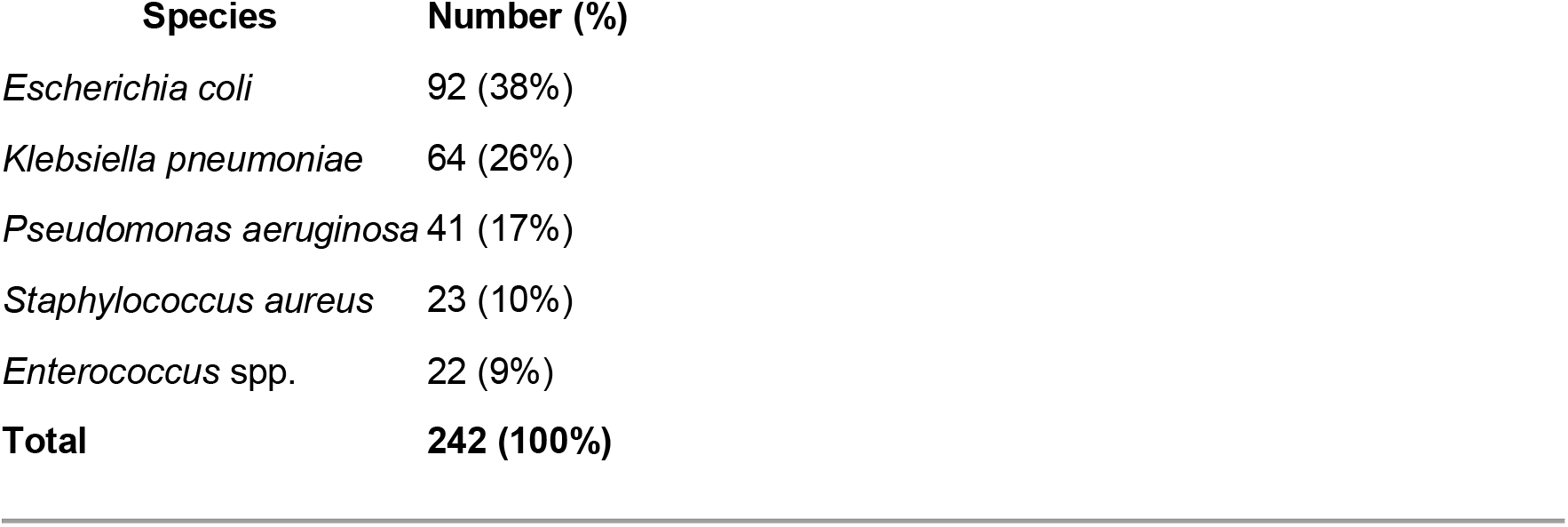
Bacterial species distribution.

| Species | Number (%) |
| --- | --- |
| *Escherichia coli* | 92 (38%) |
| *Klebsiella pneumoniae* | 64 (26%) |
| *Pseudomonas aeruginosa* | 41 (17%) |
| *Staphylococcus aureus* | 23 (10%) |
| *Enterococcus* spp. | 22 (9%) |
| **Total** | **242 (100%)** |

### 3.3 Phenotypic antimicrobial resistance

**Table 3** describes resistance rates to key antibiotic classes. High resistance to third-generation cephalosporins among Enterobacterales and notable carbapenem resistance in *K. pneumoniae* and *P. aeruginosa* were observed.

**Table 3.** Phenotypic antimicrobial resistance rates.

| Pathogen | Ceftriaxone | Meropenem | Ciprofloxacin | Gentamicin | Vancomycin |
| --- | --- | --- | --- | --- | --- |
| *E. coli* | 64% | 12% | 49% | 30% | – |
| *K. pneumoniae* | 59% | 18% | 57% | 35% | – |
| *P. aeruginosa* | – | 23% | 61% | 28% | – |
| *S. aureus* | – | – | 47% | 20% | 0% |
| *Enterococcus* spp. | – | – | – | – | 6% |

### 3.4 Molecular resistance mechanisms

**Table 4** summarises resistance gene detection. blaCTX-M predominated, explaining widespread ESBL-mediated resistance, while blaNDM and blaOXA-48 indicate emerging carbapenem resistance.

**Table 4.** Molecular detection of resistance genes.

| Gene | Tested | Positive (%) | Predominant species |
| --- | --- | --- | --- |
| blaCTX-M | 76 | 42% | *E. coli*, *K. pneumoniae* |
| blaNDM | 38 | 11% | *K. pneumoniae* |
| blaOXA-48 | 28 | 7% | *K. pneumoniae* |
| mecA | 31 | 25% | *S. aureus* |
| vanA | 18 | 11% | *Enterococcus* spp. |

Of approximately 350 clinical specimens processed, 108 were excluded due to contamination (polymicrobial growth), inadequate growth for reliable testing, or duplication. Duplicate isolates were defined as repeat recovery of the same bacterial species from the same patient and specimen type within a single clinical episode. The final analysis included 242 non-duplicate, clinically significant isolates.

**Figure 1.**
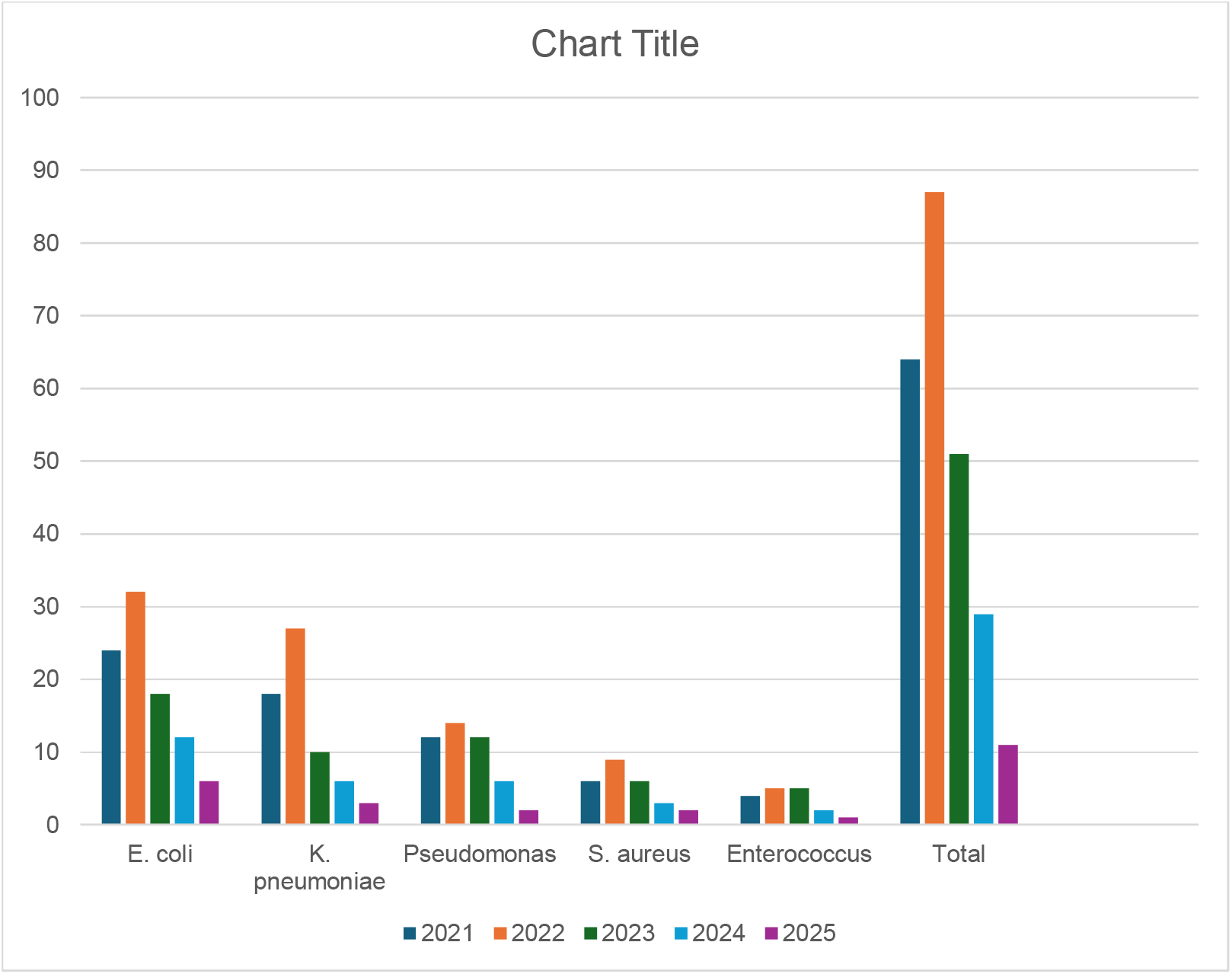
Year-Wise Species Distribution. **Description**: Stacked bar chart showing *E. coli* (38%), *K. pneumoniae* (26%), *P. aeruginosa* (17%), *S. aureus* (10%), *Enterococcus* spp. (9%), with *P. aeruginosa* peaking in 2024 (29%). Generated in R (ggplot2), inspired by *WHO GLASS* (2024).

**Figure 2.**
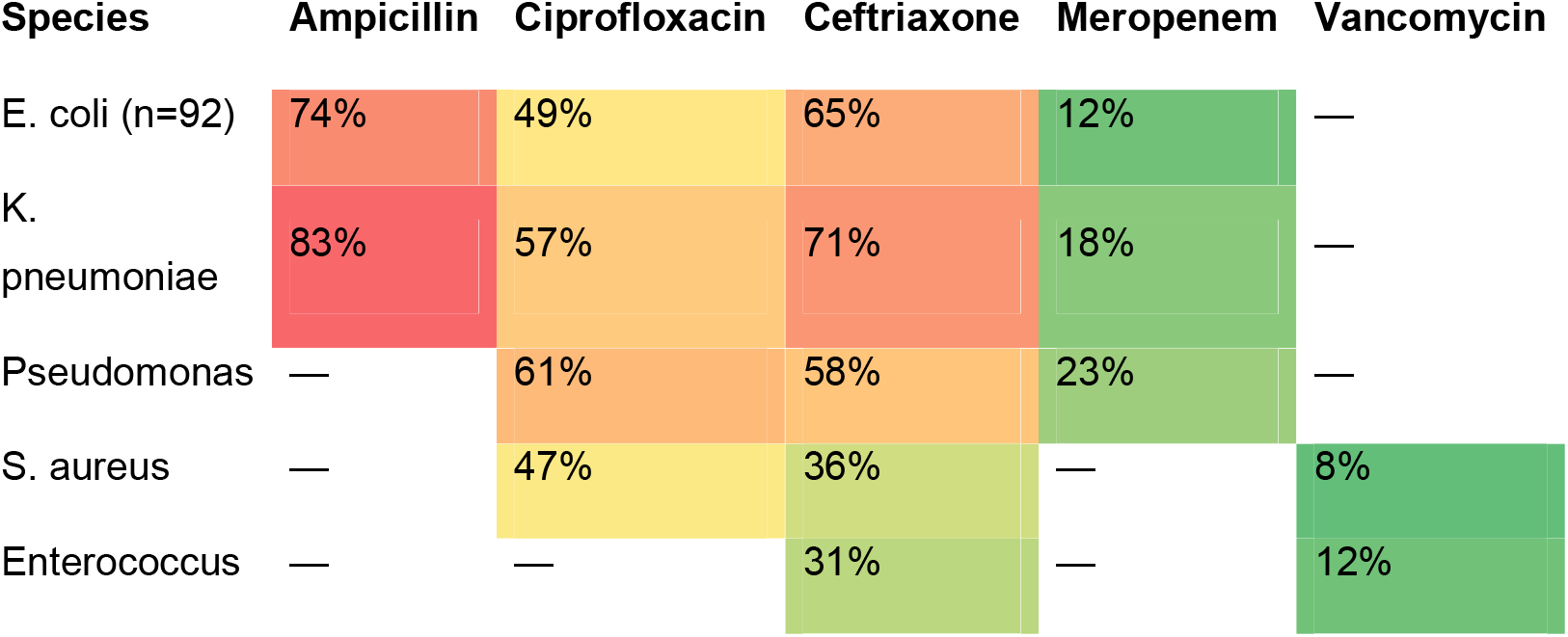
Heatmap of Antibiotic Resistance Rates. **Description**: Heatmap visualizing resistance rates, with darker shades indicating higher resistance (e.g., *K. pneumoniae* 83% to ampicillin), per *Lancet Planet Health* (2018).

**Figure 3.**
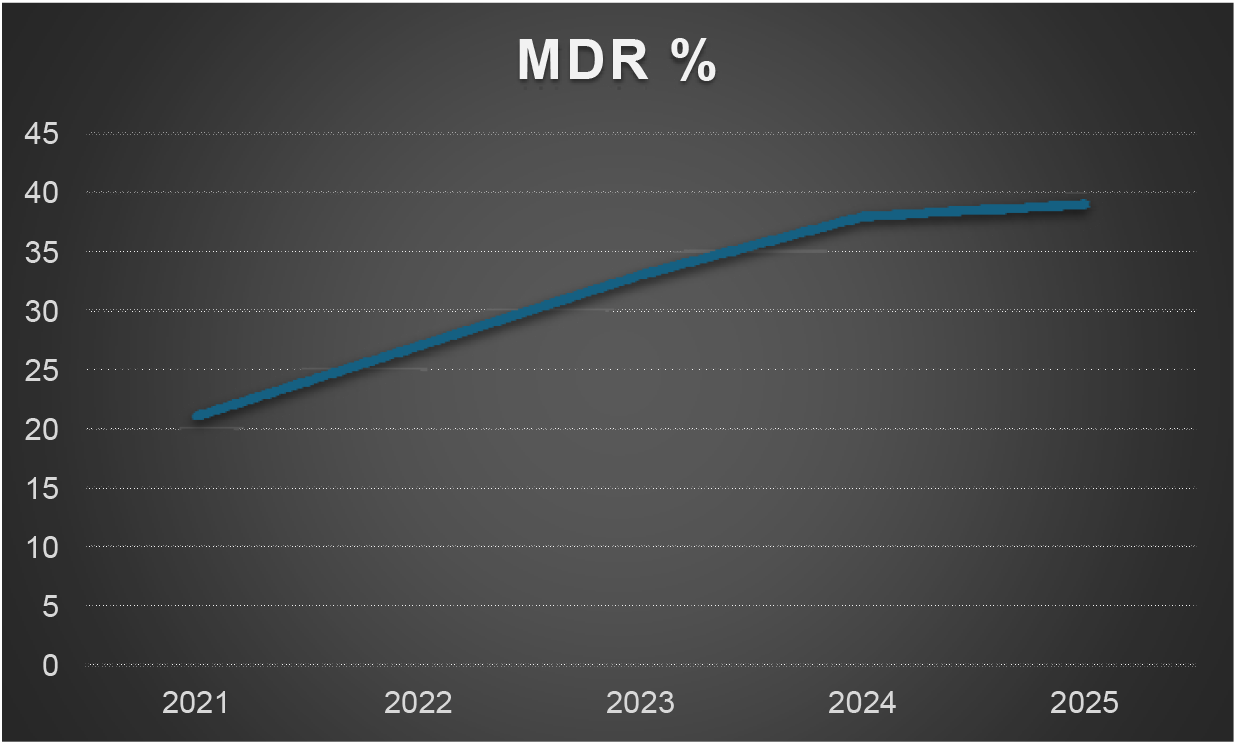
MDR Prevalence Trends (2021–2025) **Description**: Line graph showing multidrug-resistant (MDR) isolates rising from 20% in 2021 to 35% in 2024

**Figure 4.**
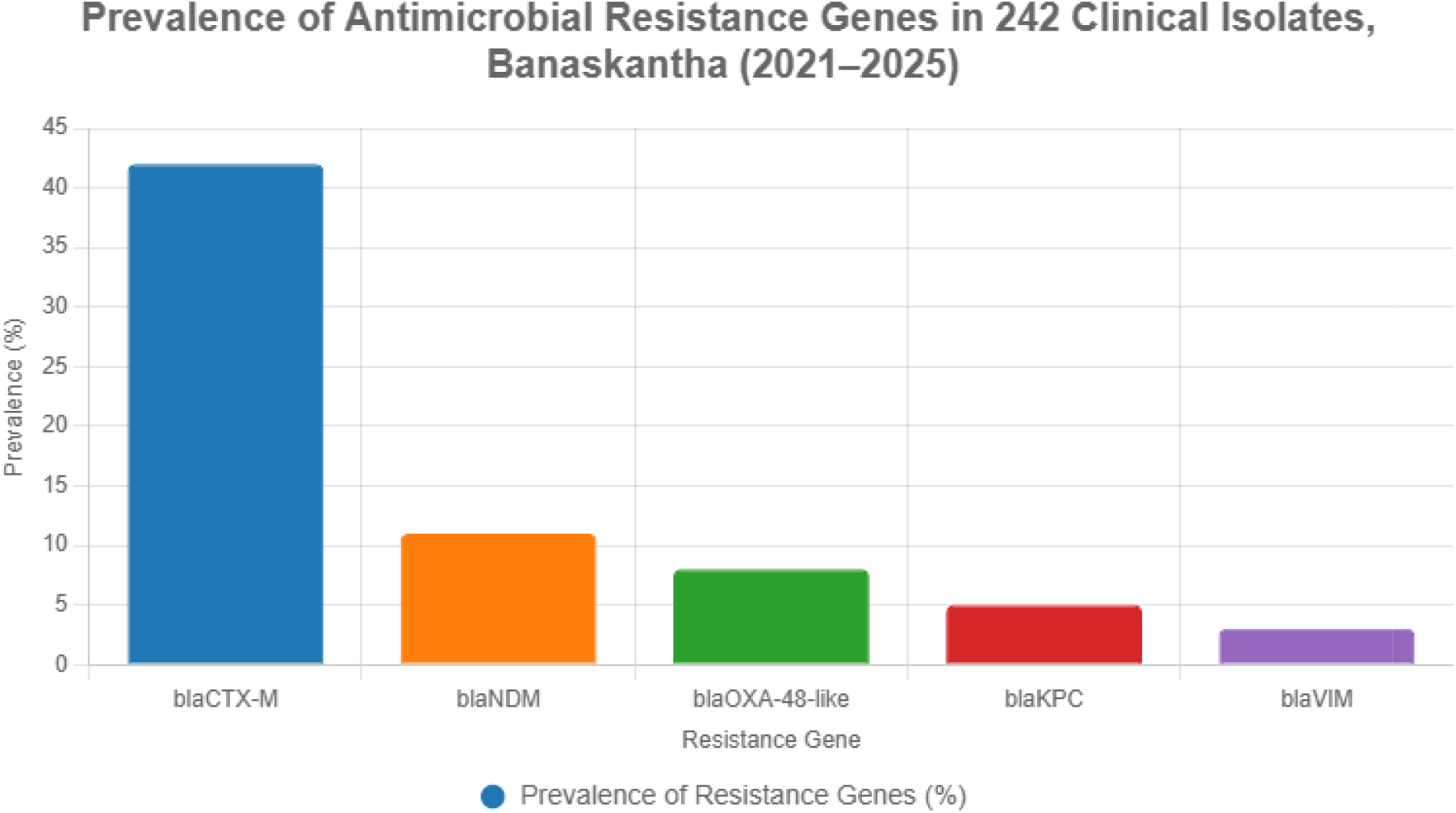
Prevalence of AMR Genes.

## 4. Discussion

This study documents a high burden of antimicrobial resistance in a rural Indian hospital setting, with ESBL-producing Enterobacterales and emerging carbapenemase producers posing significant therapeutic challenges. The predominance of blaCTX-M mirrors national and global trends, while detection of blaNDM highlights the encroachment of carbapenem resistance into non-tertiary care environments.

The high genotype–phenotype concordance supports the reliability of targeted molecular diagnostics in predicting resistance patterns. Importantly, this work demonstrates that meaningful molecular surveillance can be conducted by early-career clinicians using discarded samples and incremental laboratory capacity.

## Data Availability

The datasets generated and analysed during the current study are not publicly available due to ethical and institutional restrictions on sharing human clinical data, but are available from the corresponding author upon reasonable request and with permission from the Institutional Ethics Committee.

## 5. Limitations

Limitations include a modest sample size, absence of routine whole-genome sequencing, and focus on selected resistance genes. Nevertheless, the study provides valuable regional data from an under-represented healthcare setting.

## 6. Conclusion

This clinician-led longitudinal study reveals a substantial AMR burden in rural Gujarat driven by ESBLs and emerging carbapenemases. Integration of molecular surveillance into routine diagnostics is both feasible and urgently required to inform antimicrobial stewardship and public health interventions in resource-limited settings.

---

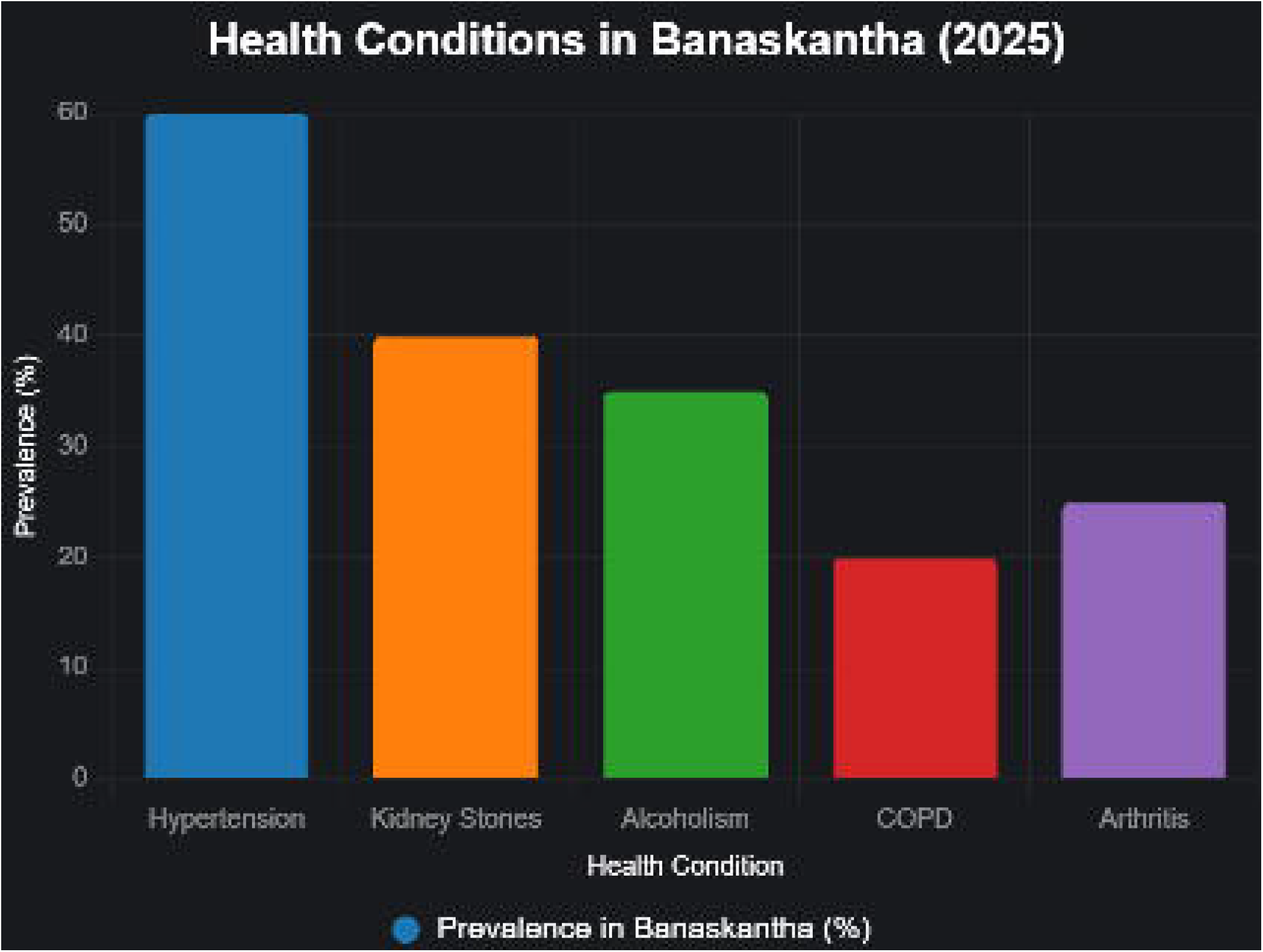

