## Supplementary File for "Molecular Surveillance of Antimicrobial Resistance in Human Clinical Isolates: A Clinician–Scientist Growth Journey from Rural Gujarat, India"

### Author Contributions

• **Conceptualization and study design:**
Dr. Atul Amarshibhai Devganiya developed the study idea based on his clinical and academic work in rural healthcare settings and designed the retrospective study combining phenotypic and molecular AMR analysis.

• **Clinical coordination and sample selection:**
The author coordinated with the hospital microbiology laboratory and selected bacterial isolates obtained during routine diagnostic work, applying clear criteria to include only non-duplicate and clinically relevant samples.

• **Microbiological analysis:**
Dr. Devganiya performed and supervised antimicrobial susceptibility testing using disk diffusion and broth microdilution methods in line with CLSI guidelines, including routine quality control procedures.

• **Molecular investigations:**
The author carried out molecular detection of key antimicrobial resistance genes (blaCTX-M, blaNDM, blaOXA-48, mecA, and vanA) using PCR techniques, with confirmation of results by Sanger sequencing.

• **Data curation and management:**
All laboratory and resistance data were carefully recorded, checked, and organised by the author to ensure accuracy, consistency, and reliability for analysis.

• **Statistical analysis and interpretation:**
Dr. Devganiya conducted descriptive statistical analyses, examined resistance trends over time, and assessed agreement between phenotypic and molecular findings, interpreting results in relation to existing AMR literature.

• **Visualization and tabulation:**
The author prepared all tables, figures, and graphical summaries to clearly present resistance patterns, molecular results, and longitudinal trends according to journal requirements.

• **Manuscript drafting:**
Dr. Devganiya wrote the complete manuscript, including all sections from abstract to conclusion, ensuring clear presentation and consistency with the journal's scope.

• **Critical revision and final approval:**
The author reviewed and revised the manuscript for scientific accuracy and clarity, approved the final version for submission, and takes full responsibility for the content of the work.
